# Diet-derived fruit and vegetable metabolites suggest sex-specific mechanisms conferring protection against osteoporosis in humans

**DOI:** 10.1101/19003848

**Authors:** Kelsey M. Mangano, Sabrina E. Noel, Chao-Qiang Lai, Jacob J. Christensen, Jose M. Ordovas, Bess Dawson-Hughes, Katherine L. Tucker, Laurence D. Parnell

**Author notes:** Corresponding authors: Laurence D. Parnell, PhD, USDA, Agricultural Research Service, JM-USDA Human Nutrition Research Center on Aging at Tufts University, 711 Washington St, Boston, MA 02111, Kelsey M. Mangano, PhD, RD, Department of Biomedical and Nutritional Sciences, University of Massachusetts, Lowell, 3 Solomont Way, Lowell, MA 01854. **ClinicalTrials.gov** Identifier: NCT01231958.

## Abstract

The impact of nutrition on the metabolic profile of osteoporosis is incompletely characterized. The objective of this cross-sectional study was to detangle the association of fruit and vegetable (FV) intakes with osteoporosis prevalence. Dietary, anthropometric and blood plasma metabolite data were examined from the Boston Puerto Rican Osteoporosis Study, a cohort of 600 individuals (age 46-79yr). High FV intake was protective against osteoporosis prevalence. Associations of 525 plasma metabolites were assessed with fruit and vegetable intake, and separately with osteoporosis status. Several biological processes affiliated with the FV-associating metabolites, including caffeine metabolism, carnitines and fatty acids, and glycerophospholipids. For osteoporosis-associated metabolites, important processes were steroid hormone biosynthesis in women, and branched-chain amino acid metabolism in men. In all instances, the metabolite patterns differed greatly between sexes, arguing for a stratified nutrition approach in recommending FV intakes to improve bone health. Factors derived from principal components analysis of the FV intakes were correlated with the osteoporosis-associated metabolites, with high intake of dark leafy greens and berries/melons appearing protective in both sexes. These data warrant investigation into whether increasing intakes of dark leafy greens, berries and melons causally affect bone turnover and BMD among adults at risk for osteoporosis via sex-specific metabolic pathways, and how gene-diet interactions alter these sex-specific metabolomic-osteoporosis links.

---

Osteoporosis (OS) is an important issue for our aging population and has emerged as a chief public health problem among aging Puerto Rican adults living on the U.S. mainland ^1^. Traditionally, OS was not viewed as a health priority for this population, yet a recent study found that 8.6% of Puerto Rican men from the Boston Puerto Rican Osteoporosis Study (BPROS) presented with OS (age-adjusted) compared with lower prevalence among non-Hispanic white men (2.3%) based on national data ^1^. Puerto Rican women showed similar age-adjusted prevalence of OS as non-Hispanic white women (10.7% vs. 10.1%). This was surprising as OS was once thought to be a condition experienced more frequently by white women than Latinas. Sex differences in the pathogenesis, prevalence and treatment of OS manifest from variations in peak bone mass and maturation, rate of annual bone loss, and disease screening methods ^2^. OS is a concern for men and women, as it typically remains undetected until a fracture occurs, which dramatically increases risk of morbidity, institutionalization and mortality ^3-5^. Although medications are available to prevent or treat bone loss, and ultimately prevent fracture, adherence is generally poor because of factors such as absence of OS symptoms, and high cost and potential side effects of medication use ^6-8^. Thus, identifying and understanding tailored, modifiable health behaviors as targets for prevention of OS are key for improving quality of life among aging adults.

Diet is one such central modifiable risk factor for OS ^9,10^. Higher fruit and vegetable (FV) intake, recommended as part of a healthy diet, has been associated with better bone health in several cross-sectional ^11-14^ and prospective epidemiologic studies ^14-16^, but not all ^17^. Across many ethnic groups, higher FV intake related to lower incidence of fracture in Chinese ^11^, European ^15^ and American ^18^ populations. Results from randomized controlled trials have been inconsistent on the effects of FV intake and bone turnover markers, ^19,20^ possibly due to poor compliance or assessment only in men and women with adequate intakes of FV. Moreover, there is increasing awareness that diet may affect subpopulations differently, warranting examination of any diet-disease interactions with a tailored nutrition approach. Thus, while there is evidence that FV intake is vital for bone health, the sex differences, biological processes and pathways underpinning this relationship remain incompletely identified.

Several mechanisms have been proposed for the effects of FV on bone health, including reduction of oxidative stress and inflammation, which in turn, reduces bone resorption and increases bone mass and strength ^21,22^. Molecular alterations central to bone health may not only inform mechanisms for the development of the disease but also may pinpoint therapeutic targets to prevent OS. Diet-derived compounds linked to bone health are not consumed as single nutrients, but rather occur as mixtures within the food matrix. Thus, interpretation of studies on individual nutrients or biochemical compounds as they relate to bone health are limited as they lack generalizability to typical dietary intakes. Therefore, the overall objective of this work was to examine associations between FV intakes and metabolomic profiles among Puerto Rican adults with and without OS. A secondary objective was to characterize the sex-specific aspects to this diet-disease association using a stratified nutrition approach.

## Results

A total of 600 men and women from the BPROS had complete dietary, metabolome and OS data (**Table 1**). For the total sample, average age was 60.0 ± 7.5 y; 70% were women; 10% had OS (11% of women, 7% of men); and average FV intake, excluding fruit juice, for the total sample was 3.0 ± 1.8 servings per day, range: (0.3-13.0).

**Table 1.** Descriptive characteristics of participants (n=600) from the Boston Puerto Rican Osteoporosis Study.

| Characteristic | Men (n=174) |  | Women (n=426) |  |
| --- | --- | --- | --- | --- |
|  | Mean or n (%) | SD | Mean or % | SD |
| Age (y) (range: 46-79) | 59.8 | 7.8 | 60.3 | 7.3 |
| BMI (kg/m<sup>2</sup>) | 30.1 | 5.1 | 33.1 | 7.0 |
| Serum 25(OH) D (nmol/L) | 18.2 | 7.0 | 19.8 | 7.5 |
| Physical activity score<sup>a</sup> | 33.1 | 6.5 | 30.9 | 3.6 |
| Current smoker (%) | 56 (32) |  | 82 (19) |  |
| Heavy alcohol consumption (%)<sup>b</sup> | 22 (13) |  | 20 (5) |  |
| Education |  |  |  |  |
| ≤8<sup>th</sup> grade | 73 (42) |  | 214 (50) |  |
| >8<sup>th</sup> grade – high school diploma | 79 (46) |  | 143 (34) |  |
| Some college | 21 (12) |  | 69 (16) |  |
| Total energy intake (kcal/d) | 2375 | 828 | 2017 | 851 |
| Total calcium intake (mg/d) | 991 | 535 | 1017 | 566 |
| Total fruit & vegetable intake (servings/d)<sup>c</sup> | 2.7 | 1.8 | 3.1 | 1.8 |
| Osteoporosis (yes, %)<sup>d</sup> | 14 (8) |  | 42 (10) |  |
| Bone mineral density (g/cm<sup>2</sup>) |  |  |  |  |
| Lumbar Spine (L2-L4) | 1.218 | 0.189 | 1.133 | 0.171 |
| Femoral Neck | 1.007 | 0.147 | 0.916 | 0.136 |

<sup>a</sup> Physical activity score: a weighted 24-h score of typical daily activity, based on hours spend doing heavy, moderate, light or sedentary activity.
<sup>b</sup> Heavy alcohol consumption defined as >2 drinks per day for men; or >1 drink per day for women.
<sup>c</sup> Total fruit and vegetable intake, excluding starchy vegetables (potatoes, cassava, plantains) and fruit juice.
<sup>d</sup> Osteoporosis at the femoral neck or the lumbar spine was defined as T-scores ≤ 2.5 (2.5 SD or more below peak bone mass).

### FV intake related to OS status

Total FV intake (servings per day) was negatively associated with prevalence of OS (P=0.03) and remained significant after inclusion of FV variety (Odds Ratio = 0.72; 95% CI = 0.56, 0.93; P=0.01); individuals without OS consumed 3.1 ± 1.8 servings FV per day while those with OS, consumed 2.3 ± 1.4 servings/day. The variety of FV comprising overall FV intake was not statistically significantly associated with OS prevalence, when controlling for total FV intake (P=0.28). FV intakes for women without and with OS were 3.2 ± 1.8 and 2.4 ± 1.4 servings per day, respectively. FV intakes for men without and with OS were 2.8 ± 1.8 and 2.0 ± 1.2, respectively.

### Metabolites significantly related to FV intake differ by sex

Metabolomics of plasma samples from 635 individuals for whom complete dietary and OS outcomes were available identified 525 known metabolites that passed specific quality control measures. Among women, 66 plasma metabolites were significantly associated with FV intake (without fruit juice), after adjustment for covariates (P<0.05), and among men 38 metabolites were significantly associated with FV intake (P<0.05) (**Supplemental Table 1, Supplemental Figure 1**). The metabolite patterns of women and men differed substantially, in that 97 of 104 total significant metabolite-FV intake associations involved unique metabolites. A few metabolite classes overlapped between men and women. Seven different constituents of caffeine metabolism were significantly lower with higher FV intake in both women and men. In the carnitine/fatty acid metabolite class, 12 different molecules, including docosahexaenoate (DHA), were higher with higher FV intake in both men and women. Lastly, of 16 glycerophospholipid metabolite levels significantly associated with FV intake, 13 had positive beta values in either men or women, or both (cf. 1-linoleoylglycerol and 2-linoleoylglycerol (18:2)) (**Figure 1**).

**Figure 1.**
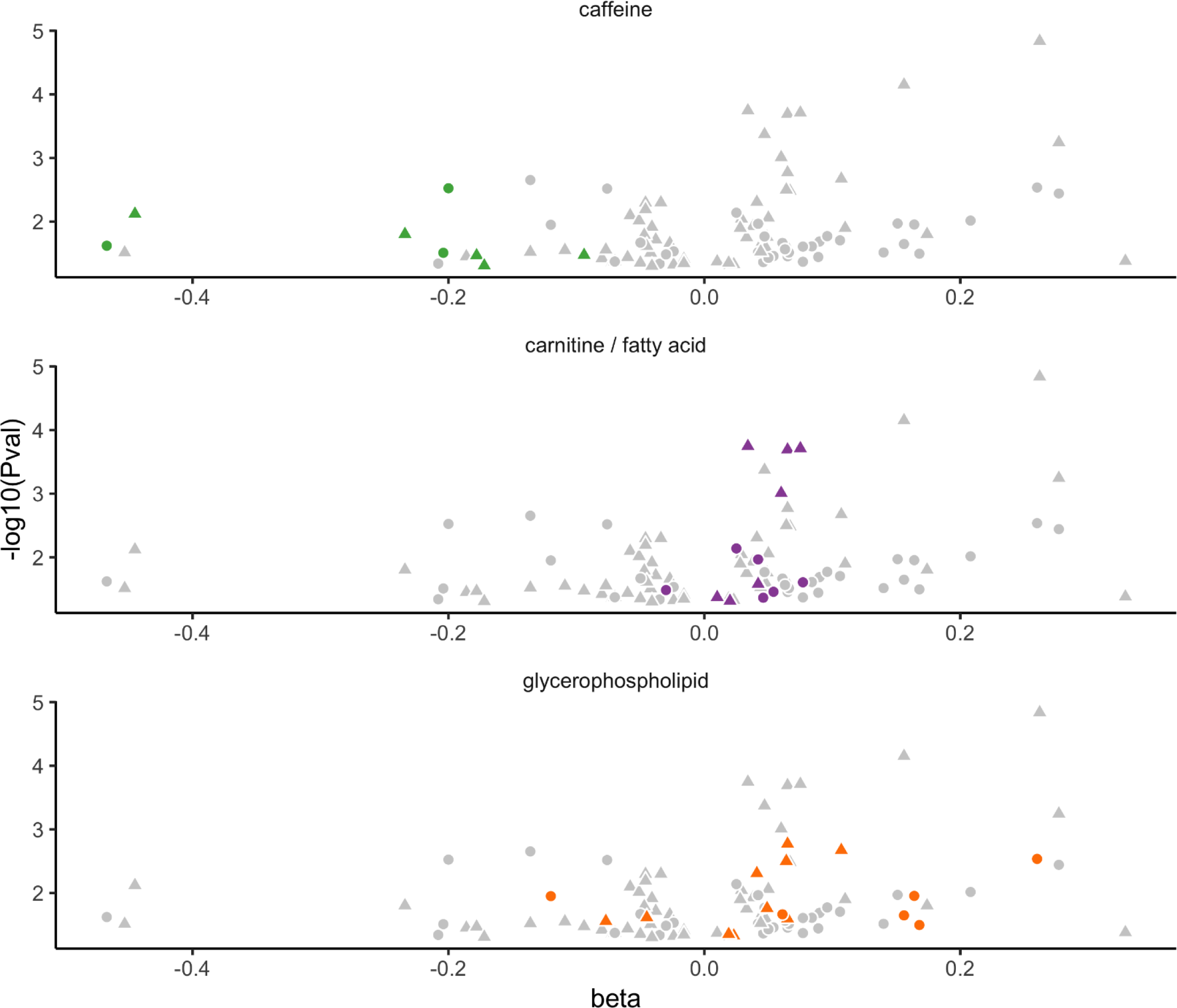
Metabolites significantly associating with fruit and vegetable intake matching specific metabolite classes. Each panel depicts a volcano plot, beta coefficient *vs* - log_10_(P-value), for all metabolites significantly associated with osteoporosis (gray) and a distinct metabolite class highlighted in color: caffeine metabolites (n=8), carnitines and fatty acids (n=13), and glycerophospholipids (n=16). Data for women are plotted with triangles, for men with circles, and are derived from **Table S1**.

Although several metabolites were associated with FV intakes in men and women separately, only seven metabolites were shared as statistically significantly associated with FV intake (**Supplemental Table 2**): docosahexaenoate (DHA; 22:6n3), iminodiacetate, homoarginine, theobromine, 1-linoleoylglycerol (18:2), 2-linoleoylglycerol (18:2), and S-1-pyrroline-5-carboxylate. Of these seven metabolites, all but homoarginine showed directionally identical beta coefficients in both men and women. Interestingly, for five of these seven metabolites, the beta coefficient was higher in men compared to women (**Figure 2**).

**Figure 2.**
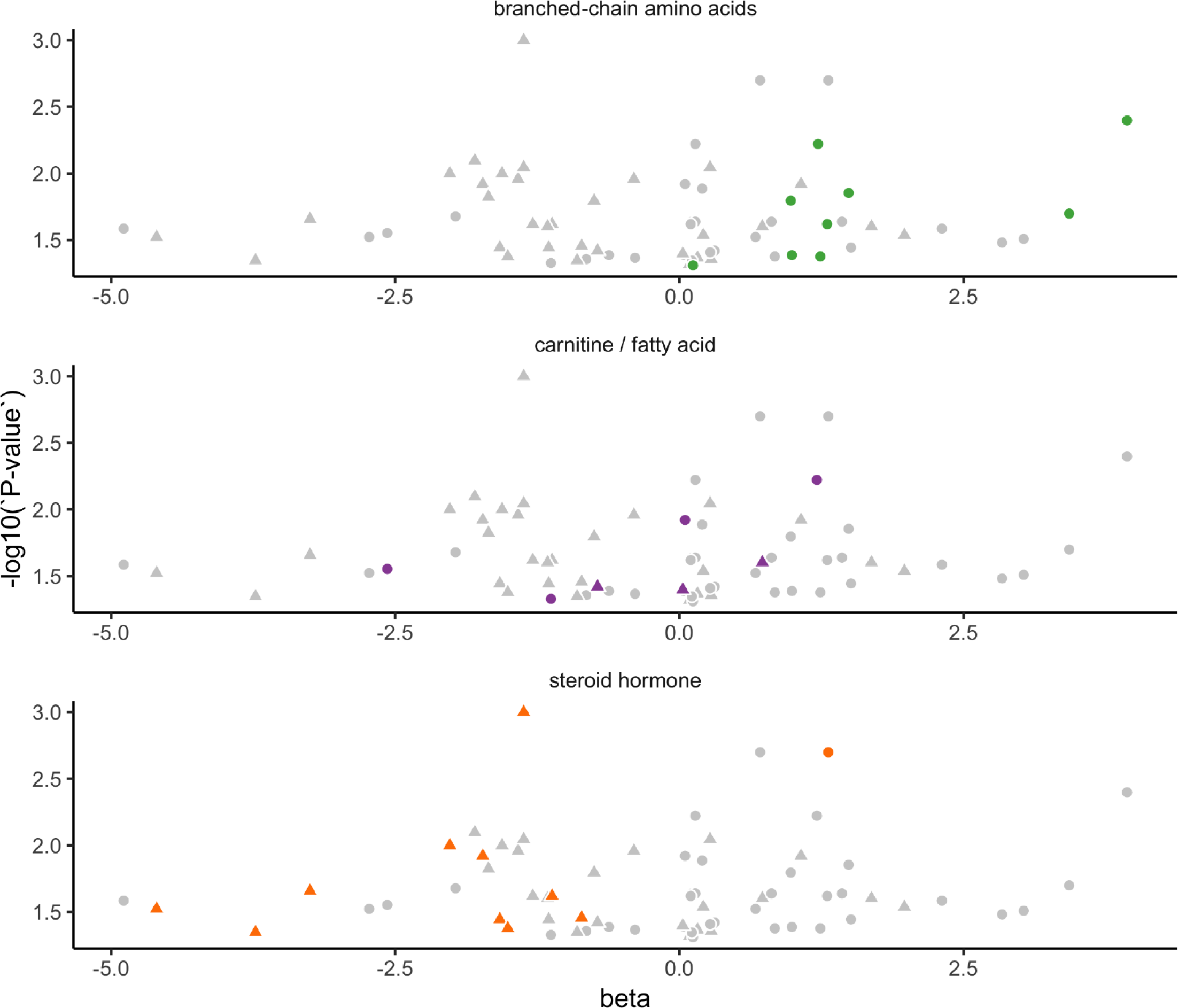
Osteoporosis-associated metabolites corresponding to specific metabolite classes. Volcano plots of beta coefficients *vs* -log_10_(P-value) depict all significantly associated metabolites (gray) with each specific metabolite class highlighted in color: branched-chain amino acids, carnitines and fatty acids, and steroid hormone metabolites. Data for women are plotted with triangles, for men with circles, and are derived from **Table S5**.

### FV-associated metabolites and enriched pathways

Pathway enrichment analysis, with FDR correction, was performed on the datasets of plasma metabolites significantly correlated with FV intake in women and men individually, to identify bioprocesses and functional modules relevant to how this food group may support reduced incidence of OS (**Supplemental Tables 3** and **4**). Notable pathways include amino acid biosynthesis, and caffeine, arginine and proline, and glycerophospholipid metabolic pathways (observed in both sexes); prostaglandin/leukotriene and α-linolenic acid/linoleic acid metabolic pathways along with biosynthesis of unsaturated fatty acids (women); and glutamate metabolism (men). Although satiety is not a defined pathway, BPROS women with high FV intakes had higher linoleoyl ethanolamide and oleoyl ethanolamide, metabolites that correlated with satiety ^23^.

### Plasma metabolites associated with OS differ by sex

Forty plasma metabolites differed significantly among men by OS status, and in women that number was 33 metabolites (**Supplemental Table 5, Supplemental Figure 2**). Similar to the metabolite sets distinguishing FV intakes, these OS-associated metabolite sets are distinct, as just two were identified as significant in both men and women with OS. N-(2-furoyl)glycine, a product of high-temperature cooking or compromised fatty acid beta-oxidation ^24^, was higher in both women (P = 0.041, beta = 0.038, beta SE = 0.019, OR = 1.04) and men (P = 0.012, beta = 0.053, beta SE = 0.021, OR = 1.05) with OS. Androstenediol (3β,17β) monosulfate, a metabolite of steroid hormone metabolism, specifically a precursor of testosterone, was lower in women with, vs. without, OS (p = 0.012, beta = -1.72, beta SE = 0.80, OR = 0.18) but higher in men with, vs. without, OS (p = 0.0027, beta = 1.32, beta SE = 0.45, OR = 3.73).

Several sex-specific metabolites merit attention. In women, ten steroid hormone metabolites were significantly lower in those with, vs. without, OS; four in the phytochemical class, and three purine/pyrimidine metabolites. In men, nine different molecules linked to branched-chain amino acid metabolism were higher in those with, vs. without, OS. In addition, six different molecules belonging to the bio-toxin/drug class were associated with OS, including three related to ibuprofen. Both women and men also had a number of significant associations between OS status and various carnitines/fatty acids, glycerophospholipids, and lysine/nitric oxide metabolites (**Figure 3, Supplemental Figure 2**).

**Figure 3.**
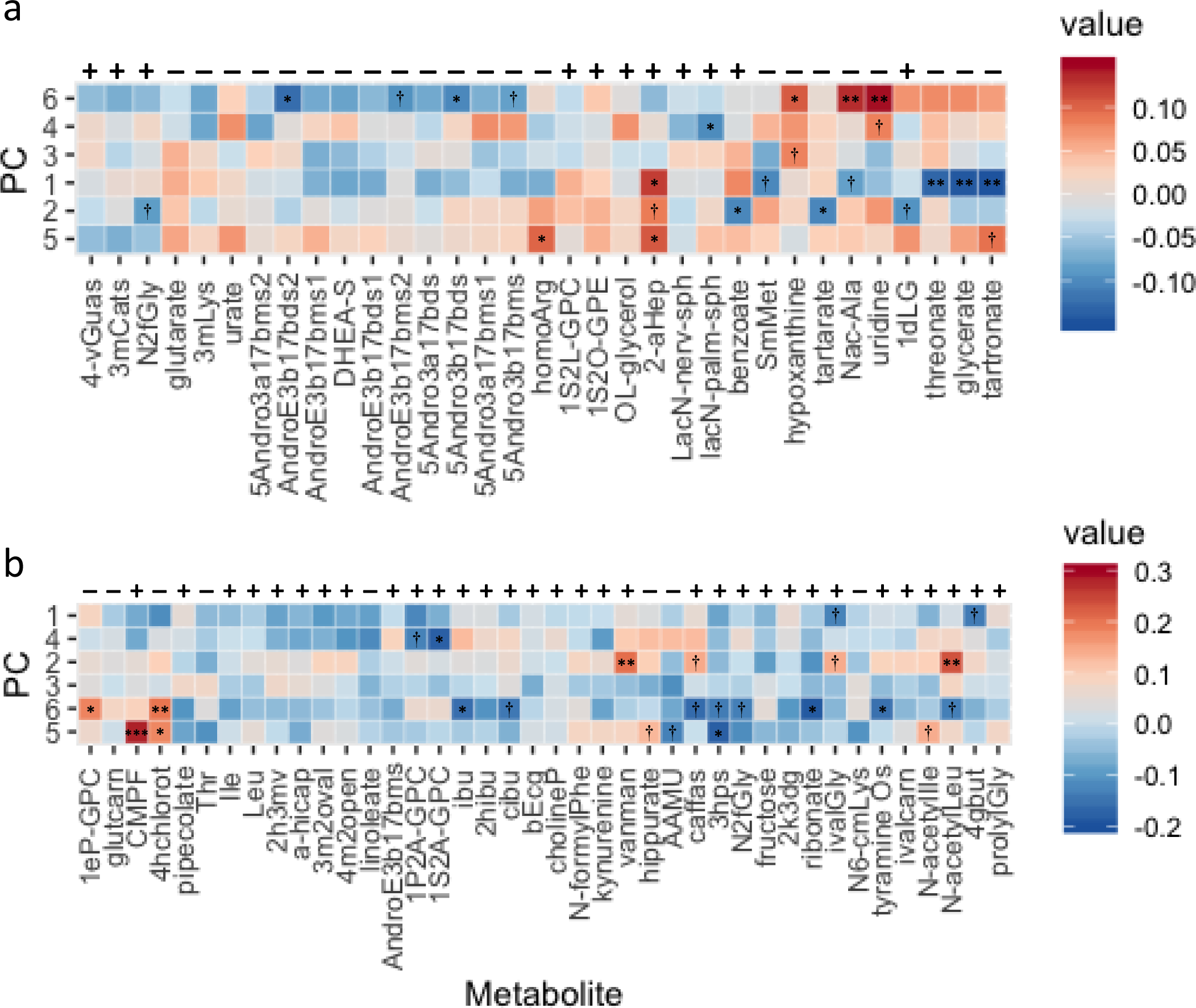
Heat maps depicting the Spearman’s rho rank correlations between osteoporosis [OS]-associated metabolites and principal component factors [PC] for fruit and vegetable [FV] intake. Above each heat map are minus / plus symbols indicating the direction of association (beta) of that metabolite with OS, panel a) for women and panel b) for men. The factors for FV intake and the metabolites are both ordered according to hierarchical clustering based on *hclust* in R. Individual panels are annotated with asterisks (**\***) for those metabolites whose correlation with the factor reached statistical significance, or a † when significance of that correlation was <0.10. Significance of the correlation at P<0.01 and P<0.001 are indicated by ** and *** respectively. Panels where the color representing the correlation are opposite to the beta of association with OS can be considered protective to some extent. Abbreviations for metabolites are defined in **Table S9**.

Comparing the metabolites that discriminated FV intake (n=66) and those that characterized OS status (n=33) there was an indication of five shared molecules in women: glutarate, glycerate, homoarginine, tartarate and tartronate, all of which were higher with higher FV intake, and lower with OS (compared to those without OS). For men, there was no overlap of metabolites associated with FV intake and OS status.

### OS metabolites and enriched pathways

Pathway enrichment analysis was performed separately on the datasets of plasma metabolites significantly correlated with OS for women and men to identify relevant bioprocesses and functional modules (**Supplemental Tables 6** and **7)**. For women, as noted above, ten of the 33 metabolites correlated with OS were sulfate conjugates of steroid hormones. However, many of these compounds have not been mapped to metabolic pathways used by the analysis tools chosen, hindering a thorough enrichment assessment. Nonetheless, a steroid hormone module was significant (FDR P-value 0.017). Other pathways and bioprocesses of note include phosphatidylcholine and phosphatidylethanolamine biosynthesis, adenosine/nucleotide metabolism, lysine metabolism, glyoxylate/dicarboxylate metabolism, and degenerative disc disease pathway. Results from the Mbrole tool highlight the degenerative disc disease pathway, driven by significantly lower hypoxanthine, urate and uridine with OS (**Supplemental Table 6**). These metabolites also are indicators of adenosine degradation, which was significant at FDR P=0.0296. For men, pathways of note showing enrichment based on metabolite differences between those with and without OS were BCAA metabolism, and glycerophospholipid metabolism, including conversion of phospholipid to linoleate within linoleic acid metabolism and phosphatidylcholine biosynthesis (**Supplemental Table 7**).

### Link between FV-associated metabolites and the steroid hormone biosynthesis pathway

Ten of 33 OS-associated metabolites are steroid hormone derivatives. Metabolites significantly related to high FV intake that can inhibit one or more enzymatic processes in the KEGG steroid hormone biosynthesis pathway include 1-methylnicotinamide (beta +), arachidonate (+), deoxycholate (-), naproxen (-), piperine (+) (all P<0.05), and fatty acids linoleate, oleate/vaccinate, palmitate and stearate (all P<0.097, with beta +) (**Supplemental Table 8**).

### Correlations between FV PCA factors and OS-associated metabolites

Principal components analysis of total FV intake generated six meaningful factors: 1) traditional/sofrito, 2) American vegetable, 3) tropical fruits, 4) other fruits, 5) berries and melons, and 6) dark leafy greens. Correlations between FV derived factors and OS-associated metabolites are presented as heat maps (**Figure 4**). In women, six OS-associated metabolites correlated with factor 1 (traditional/sofrito) at P<0.10, with positive correlation coefficients (metabolite by factor 1) suggesting that they are associated with both OS and higher intakes of these vegetables. This relationship in men differed, where ten OS-associated metabolites correlated with FV factor 6 at P<0.10, with correlation coefficients opposite to that for OS. Metabolites significantly associated with PC factors indicating higher intake, while also associated with less likelihood of OS included, in men: 1-(1-enyl-palmitoyl)-GPC (PC: leafy greens), 4-hydroxychlorothalonil (PCs: berries/melons & leafy greens), and hippurate (PC: berries/melons) and; in women: homoarginine (PC: berries/melons), hypoxanthine (PC: tropical fruits & leafy greens), N-acetyl-beta-alanine (PC: leafy greens), uridine (PCs: other fruits & leafy greens) and tartronate (PC: berries/melons).

**Figure 4.**
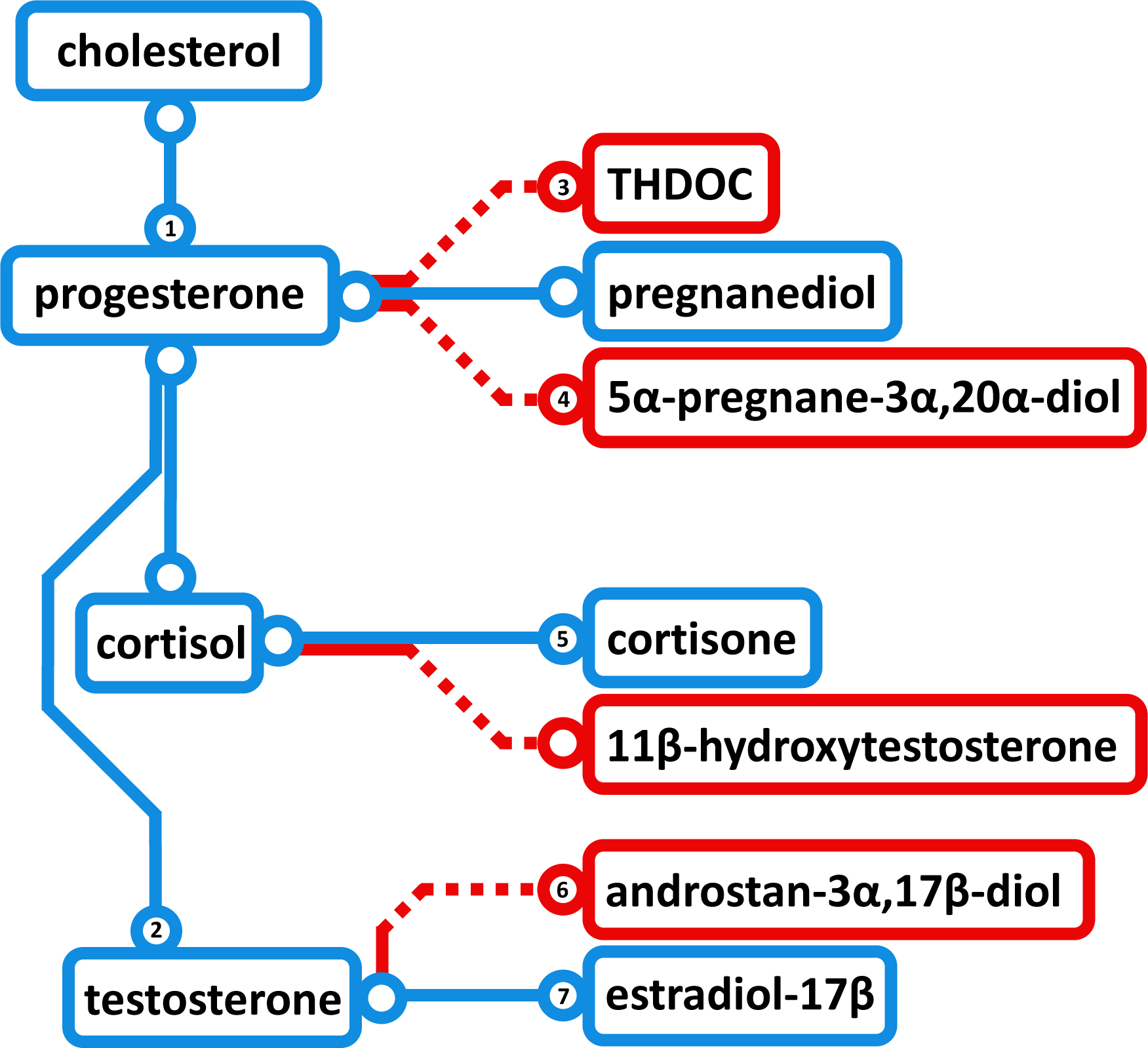
Schematic of steroid hormone biosynthesis, proceeding from cholesterol to various hormones via successive enzymatic reactions. Blue paths indicate synthesis modules predicted not to be repressed in women based on the observed osteoporosis-associated metabolite profiles. Red paths are predicted to be partially inhibited based on enzyme inhibition data mined from BRENDA, with numerals indicating those metabolite-enzyme pairs listed in **Table S8**. THDOC, tetrahydrodeoxycorticosterone (3α,21-dihydroxy-5α-pregnan-20-one).

### Correlations between OS-PCA factors and OS status

The ten steroid hormone metabolites that were negatively associated with OS in women were the main drivers of factor 1 (PC1), which accounts for 21.2% of the variance (**Supplemental Figure 3**). Age was not a significant factor (P=0.144) in predicting OS status compared to PC1 (P=0.0068, logit model). Further analysis of the PC data showed that women with low factor 1 score had higher likelihood of OS (P = 9.2E-08) (**Supplemental Figure 4**). In men, nine of the ten OS-associated metabolites that correlated well with both bone health and FV factor 6 (dark leafy greens) (**Figure 4B**) showed good separation in the PC plot generated from the OS-associated metabolites (**Supplemental Figure 5**). N-acetylleucine was the exception.

## Discussion

To our knowledge, this is the first study to characterize plasma metabolite patterns for osteoporosis in conjunction with the beneficial impact of FV intake in men and women on bone. The current study identified metabolite differences associated with greater FV intake and showed sex-specific associations to metabolic pathways in older Puerto Rican adults. Similarly, the pathways represented by the OS-associated metabolites differed by sex, suggesting that diet may exert sex-specific mechanisms to affect bone health. These results warrant investigation into potential personalized nutrition advice for bone health, explicit to FV intakes that may act upon different sex-specific metabolic pathways related to bone.

In cross-sectional studies, higher FV intake has been associated with greater BMD in Chinese adults ^11^, and non-Hispanic white men and women ^14^. Results from the CHANCES project, a large cooperative study of osteoporotic fracture, implied that FV intake below 1 serving/day increased risk of hip fracture by 39% ^18^. Analysis of a Swedish cohort of men and women showed that FV intake under 5 servings/day significantly increased hip fracture risk ^15^. In a meta-analysis, higher intake of vegetables, but not fruit, was associated with lower incidence of fracture ^25^. In the current study, greater FV intakes were significantly associated with reduced prevalence of OS among Puerto Rican older adults, but not variety. FV provide micronutrients that affect bone positively, including vitamin K, folate, magnesium, potassium, vitamin C and carotenoids ^26^. Moreover, greater FV intake may modulate the gut microbiome, thereby altering metabolic pathways related to bone ^27,28^. Identifying such metabolic pathways is required to grasp how diets rich in FV influence bone health and reduce risk of fracture.

In the current study, greater FV intake in women was associated with higher arachidonate, as well as PUFA DHA, n-3 DPA and EPA. These metabolites may decrease risk of OS by reducing low-grade chronic inflammation and supporting a shift in mesenchymal stem cell lineage commitment ^29^. PUFA also have been shown to inhibit RANKL-induced osteoclast formation in a dose-dependent manner, with arachidonic acid- and DHA-mediated inhibition the strongest effectors ^30^. Women with high FV intake were shown here to have higher homoarginine and piperine concentrations, and homoarginine deficiency has been associated with increased bone turnover ^31^, while piperine has been shown to alleviate osteoclast formation ^32^. Greater FV intake in women correlated with lower concentrations of several plasma metabolites, including deoxycholate, gamma-carboxyglutamate, kynurenine, S-1-pyrroline-5-carboxylate, S-adenosylhomocysteine and the NSAID naproxen. Lower kynurenine may protect bone, as elevated kynurenine has been linked to accelerated skeletal aging by impaired osteoblastic differentiation and increased osteoclastic resorption ^33^. Finally, higher intake of FV was associated with lower deoxycholate, gamma-carboxyglutamate, S-1-pyrroline-5-carboxylate and S-adenosylhomocysteine, metabolites with potential to benefit bone. For example, higher deoxycholic acid and S-adenosylhomocysteine were related to lower BMD ^34,35^, and gamma-carboxyglutamate to greater likelihood of OS ^36^. Overall, it is clear that several metabolites related to FV intake in this study have the potential to positively influence bone health in women.

In men, higher FV intake was associated with lower nicotinamide and S-1-pyrroline-5-carboxylate, which were reported to inhibit osteoclast differentiation ^37^ and promote osteopenia, respectively ^38^. Greater FV intake in men was associated with higher dimethylglycine (DMG) and myo-inositol. Low plasma DMG has been associated with low BMD and increased risk of hip fracture ^39^, while higher BMD has been associated with phytate (myo-inositol hexaphosphate) intake ^40^. Men with higher FV intake also had higher DHA levels, which was shown to positively influence bone health ^(46)^ potentially through inhibiting late-stage osteoclastogenesis ^41^, although, in the current study, this association was not as strong as that observed in women.

Of 12 different pathways and processes showing enriched representation by the set of 66 metabolites associated with FV intakes in women, several merit highlighting. First, biosynthesis pathways of unsaturated fatty acids (P=0.0032) and α-linolenic acid and linoleic acid metabolism (P=0.0036) indicate a relation between bone function and bone marrow (BM) fat. For example, significantly lower unsaturation of BM lipids has been noted in OS patients than in controls ^42^, with low unsaturation and high saturation lipid concentrations in diabetes patients with prevalent fractures ^43^. Second, α-linolenic acid metabolism is pertinent to the function of the G-protein-coupled receptor GPR40, a long chain unsaturated fatty acid receptor, which is regulated by estrogen and involved in *in vivo* and *in vitro* osteogenic differentiation ^44^. Third, reduced glycine degradation can positively affect creatinine biosynthesis, and at least one meta-analysis showed that creatine supplementation may improve BMD ^45^. Although many metabolites detected as significant differed between men and women, a few pathways were shared: arginine and proline metabolism, protein synthesis, and caffeine metabolism. Caffeine metabolites were lower in both men and women with greater FV intake. However, coffee intake did not differ significantly by FV intake or OS status intake; hence the observed lower caffeine metabolites may indicate altered liver function ^46^. Lower arginine and proline metabolism, as observed here for proline, S-adenosylhomocysteine and S-1-pyrroline-5-carboxylate, agree with observations indicating that dysregulation in arginine metabolism may partially explain bone related complication of diabetes ^47^. Approximately 43% of the current BPROS cohort had diabetes at the time of measurement. Protein metabolism has been shown to influence bone health positively, where amino acid-sensing mechanisms may be important in calcium metabolism and bone homeostasis ^48^. As a whole, pathways enriched by FV intake in this study may influence bone health via modulations in BM fatty acids, and regulation of amino acid metabolism.

Of 33 plasma metabolites significantly different in women with OS compared to those without, ten steroid hormone metabolites were significantly lower among women with OS. These include mono- and disulfate byproducts of dehydroepiandrosterone (DHEA-S) and testosterone. Among men, only androstenediol (3β,17β) monosulfate differed significantly by OS status. It is well established that estrogens and androgens slow the rate of bone remodeling and protect against bone loss ^49,50^. Conversely, estrogen loss leads to increased remodeling and negatively affects the balance between bone resorption and formation in favor of resorption ^51^. The current study corroborates that loss of steroid hormones with age, particularly among women, is detrimental for bone and increases prevalence of OS.

Three different compounds positively associated with FV intake in women might partially inhibit ten enzymes and five different reactions in steroid hormone biosynthesis. Another four metabolites with trending associations with FV intake might contribute to inhibiting these same five reactions. Additionally, another three metabolites (one trending) showed negative association with FV intake and potential for reduced inhibition of other enzymes and outputs of this pathway. We interpret these data to suggest that a diet high in FV in women changes the levels of specific metabolites that can alter the flux through the steroid hormone biosynthesis pathway, promoting progesterone, testosterone and estradiol-17β production (**Figure 5**, Supplemental Table 8). Detailed analysis of OS-associated metabolites supports this interpretation, in that ten different steroid hormone metabolites were negatively associated with OS prevalence in women, and that factor 1 from PCA of these 33 metabolites includes strong loadings for all ten of these OS metabolites. Importantly, age was not a significant factor in predicting OS status, in contrast to the impact of FV intake on OS status via steroid hormone biosynthesis.

Steroid 5α-reductase (SRD5A, EC 1.3.1.22), inhibited by five of the FV-associated metabolites, catalyzes dihydrotestosterone (DHT) formation, and its inhibition promotes estrogen production. Treating 51 men with recurrent or metastatic prostate cancer with a bispecific SRD5A inhibitor yielded significantly elevated estradiol ^52^. Therapeutic SRD5A inhibitors are prescribed most commonly for male pattern baldness. Due to the direct DHT reduction and because DHT is an key hormone in bone anabolism, several studies examined how chronic use of such inhibitors affects bone health in men, implying that SRD5A inhibitors demonstrate neutral impact on BMD^53-55^ with one study of 14,152 men showing reduced fracture rates over 9-yr follow-up ^56^. It was then postulated that the neutral or slightly positive effects on bone resulted from an indirect increase in circulating estrogen. Thus, habitually high intake of FV may increase estrogen levels via SRD5A inhibition.

Upregulation of STS, steroid sulfatase (3.1.6.2), is another enzyme positively influenced by higher FV intakes. Post-menopausal women have substantially lower gonadal estrogens in circulation, resulting in altered bone remodeling, decreased bone density and increased OS risk ^57,58^. Yet, postmenopausal women tend to have high circulating sulfated steroids, particularly estrone sulfate and DHEA-S ^59^. Conversion of these precursors to active estrogens by STS may provide some of the estrogen needed to maintain bone density in postmenopausal women. Preosteoblastic cells possess STS activity and can upregulate estrogen production in bone ^60^. The current study suggests that greater FV intakes may modulate STS activity and promote a healthy bone phenotype. However, current data do not clarify if reduced inhibition of STS via lower deoxycholate in women with high FV intake directly leads to greater STS production and/or activity (Supplemental Table 8). Metabolites associated with a dietary pattern that itself associates with bone health could contribute to inhibiting pathways relevant to bone phenotypes. Further research must determine if specific FV-associated metabolites affect steroid hormone biosynthesis. It is also necessary to ascertain if altered regulation of these biosynthetic enzymes by FV intakes affects bone health via systemic changes in concentration and/or via regulation of sex steroid hormones directly within bone cells.

In addition to steroid hormone metabolites, differences in several other compounds are notable among women with, vs. without, OS. Higher ceramides lactosyl-N-nervonoyl-sphingosine and lactosyl-N-palmitoyl-sphingosine could be indicators of osteoblast cell death via caspase-9-mediated apoptosis ^61^. Lower threonate in OS women could relate to tissue mineralization and bone resorption ^62,63^. Lower homoarginine is an indicator of higher bone turnover ^31^. Glutarate stimulates SLC22A8/Roct organic anion transport activity, and lower glutarate in OS women may compromise the excretion of Roct substrates such as bone turnover factors ^64^.

Men with OS presented with differences in several compounds relevant to this disease. Higher ibuprofen and its metabolites carboxyibuprofen and 2-hydroxyibuprofen suggest the possible occurrence of joint pain. Hippurate, lower in men with OS, stimulated bone-forming gene expression in mice ^65^. Higher plasma ribonate and kynurenine may indicate chronic kidney disease (CKD) ^66^. Rats with CKD were shown to have elevated peripheral kynurenine and pathological changes in bone structure ^67^. Here, plasma N6-carboxymethyllysine (N6-cmLys), a major product of non-enzymatic glycosylation of collagen by glucuronic acid, was lower in OS men ^68^, in contrast to literature showing higher N6-cmLys in adults with OS ^69^. However, it is not clear whether glycosylated end products, such as N6-cmLys are a causal phenomenon of OS or an epiphenomenon. Arguably, the most impressive of the OS-associated metabolites in men in this study, is the set of nine that function in BCAA metabolism, including isoleucine, leucine and seven affiliated metabolites. In men with, vs without, OS, all nine were higher. Although, to our knowledge, our study is the first to link elevated BCAA metabolism with OS, others have proposed a role for altered BCAA metabolism in osteoarthritis ^70,71^, and these metabolites may prove useful as biomarkers of the combined effects of metabolic dysfunction and disease progression.

In this analysis, FV intake and OS status metabolite patterns differed greatly by sex. Greater FV intake may affect different mechanisms as bone health declines, perhaps because high FV intake, although providing many benefits to bone ^14^, is itself a good marker of an overall healthy dietary pattern ^72^. Hence, it is not surprising that so few OS-associated metabolites correlate with FV PC factors at P<0.05. PCA produced six major factors representing FV intakes within this adult population, showing that dark leafy greens and berries/melon were significantly associated with reduced likelihood of OS in both men and women. Although to our knowledge there is little to no literature examining the relation between dark leafy greens, berries and bone health, dark leafy greens are the primary source of vitamin K in the diet, which has been shown to lower risk of fracture ^73,74^ and are high in magnesium, which recently was shown to be important for bone health in this cohort ^75^. In addition, berries are an excellent source of vitamin C and carotenoids, which have been associated independently with higher BMD ^76^ and lower risk of fracture ^77^. Here, the association between the dark leafy green factor and reduced risk of OS was stronger in men, where dark leafy green intakes were negatively correlated with ten of the 40 OS-associated metabolites, while in women, they correlated with three.

This study had several strengths. To date, this is one of the largest studies of bone health among Puerto Rican adults living on the US mainland, although there was a small sample of men with osteoporosis. A comprehensive analysis of metabolomics was performed on a large sample of community dwelling older adults. In addition, bone measures and assessment of osteoporosis were completed using the benchmark (DXA) and standard definitions for the disease. Limitations to the current study include the cross-sectional design and inability to assess causality. The use of the FFQ has inherent limitations because of the lack of detailed information on portion sizes and specific recipes with the potential for systematic errors that could have resulted from under- or over-reporting of food intakes. As was used in the current study, energy adjustment can partially mitigate systematic errors. The FFQ is best suited for ranking typical food group intakes as were used in this study.

In summary, women with greater FV intakes had higher concentrations of metabolites known to inhibit specific branches of the steroid biosynthesis pathway, enhancing greater estrogen production. In both men and women, a dark leafy green dietary factor and a berries/melon factor were significantly related to reduced likelihood of OS. These data warrant future investigation into whether increasing FV intake, particularly dark leafy greens and/or berries/melons, may causally affect bone turnover and BMD among adults at risk for osteoporosis. In addition, future research is warranted to investigate tailored nutrition interventions that may differentially alter bone health by sex.

## Methods

### Study population

This study included data for 600 participants aged 46-79yr from the Boston Puerto Rican Osteoporosis Study (BPROS), an ancillary study to the Boston Puerto Rican Health Study. The parent BPRHS cohort included 1504 Puerto Rican adults aged 45-75yr recruited from the Greater Boston Area through door-to-door enumeration and community-engaged activities ^78^. Of 1504 Puerto Rican adults who completed the baseline interview, 1267 participated in a 2-yr follow up visit. All who completed the 2-yr visit were invited to join the BPROS. A total of 973 participants were re-consented for the BPROS; 205 declined participation, 13 had moved from the area, 47 had difficulty scheduling the interview, 11 were lost to follow up, two did not participate for other reasons, and 20 had died since the 2-yr interview. Four participants did not complete the 2-yr interview but were re-consented for BPROS. Those who declined to participate were older (60.9yr vs. 58.7yr, *P*<0.001) and more likely to have type 2 diabetes (47.8% vs. 40.4%, *P*=0.03) than those who participated in the BPROS (no differences were noted by sex, *P*=0.91; smoking status, *P*=0.16; physical activity score, *P*=0.42; or activities of daily living, *P*=0.34). BPROS participants were invited to the Bone Metabolism Laboratory at the Jean Mayer USDA Human Nutrition Research Center on Aging at Tufts University to complete an in-person interview, body composition and BMD measures with a trained, bilingual interviewer. Of 973 participants consented for BPROS, 21 participants were removed from analyses due to invalid BMD measurements at the lumbar spine and/or femoral neck. 817 BPRHS parent cohort samples were sent to Metabolon, Inc (Morrisville, NC USA) for metabolomic analysis. With the final data merge, 635 participants presented with diet, bone and metabolomics data; 35 of these participants were missing one or more FV food groups and/or their diet records were deemed invalid. Thus, the total sample size in current analyses included data from 600 BPROS participants. All participants provided written informed consent. The study was approved by the IRBs at Tufts University and the University of Massachusetts, Lowell, and adhered to the ethical principles of the declaration of Helsinki.

### Data Collection

Study data were collected and managed using REDCap electronic data capture tools hosted at the University of Massachusetts, Lowell ^79,80^. REDCap (Research Electronic Data Capture) is a secure, web-based software platform designed to support data capture for research studies, providing 1) an intuitive interface for validated data capture; 2) audit trails for tracking data manipulation and export procedures; 3) automated export procedures for seamless data downloads to common statistical packages; and 4) procedures for data integration and interoperability with external sources.

### Measures of Bone Mineral Density and Osteoporosis

BMD was assessed with DXA on a GE-Lunar Model Prodigy scanner (GE Lunar, Madison, WI, USA), with weekly calibration using an external standard (aluminum spine phantom; Lunar Radiation Corp). As reported previously, the root mean square precision was 1.31% and 1.04% for BMD of the femoral neck (FN) and lumbar spine (LS, L2-L4), respectively ^81^. The DXA measures were completed using DXA acquisition software version 6.1 and analysis version 12.2. The right hip was scanned per standard procedures, unless the participant reported hip fracture or joint replacement in that hip. The study endocrinologist (BDH) reviewed all scans identified as having a *T*-score >4.0 for non-anatomical parts and for extraskeletal calcification, and excluded 32 participants (25 for the LS and 7 for the FN). OS at the FN or the LS was defined as *T*-scores ≤ 2.5 (2.5 SD or more below peak bone mass) ^82^. Individuals were classified as having OS if they had OS at either the hip or spine.

### Dietary Assessment

Analyses used dietary measures from the interview closest to the BMD measurement (at 2-y follow-up visit). A food frequency questionnaire (FFQ) adapted and validated for use in this population of Hispanic adults was used to assess usual dietary intake. The traditional Puerto Rican diet differs considerably from both the general US population and from other Hispanic subgroups (*e*.*g*. Mexican Americans). Hence, the food list for the FFQ was developed using the National Cancer Institute/Block food frequency format but modified with data from the Hispanic Health and Nutrition Examination Survey dietary recalls for Puerto Rican adults. For example, foods like plantains, and specific soup and rice-dish recipes, as well as appropriate portion sizes, were added to the FFQ. The BPRHS FFQ is a better estimator of dietary intakes in this Hispanic population than the original Block questionnaire ^83^. It has been validated against plasma carotenoids ^84^, vitamin E ^85^, vitamin B6 ^86^ and vitamin B12 ^87^ in Hispanic adults aged 60yr and older. Information on nutrient intakes were calculated from the FFQs using the Nutrient Data System for Research software.

Servings of FV were obtained by dividing the gram amount of each food by the reference serving amount in the USDA Food Guide Pyramid. A composite variable for total FV intakes (servings per day) was calculated as the sum of all FV intakes, excluding starchy root vegetables (potatoes, cassava, plantains), vegetables considered as fat sources (avocado and olives) and fruit juices. Variety in FV intake was defined as the total number of unique fruits and vegetables consumed at least once per month over the past 12 months. Variety score was regressed on total FV servings per day to generate residuals that were then included in models to predict OS. The use of residuals accounts for variation in types of FV consumed, independent of total FV intake, as reported ^88^.

To characterize the common groupings of FV consumed by this cohort of Puerto Rican adults, we performed principal components analysis (PCA) on the intakes of 42 FV items, which was accomplished in several steps. First, FV were condensed to 42 predefined groups, based on nutrient-composition similarities. Only foods included in the total FV intake calculation were included in the food groupings. Each food group was then calculated as a percent contribution to total FV servings per day (food group x1 = x1 servings intake per day/total FV servings per day * 100). Because PCA is sensitive to outliers, data were evaluated to ensure that no participants with FV contributions from food groups more than 0.5 SD beyond the mean intake contribution for that group were included (2 participants excluded). PCA factors were generated using the PROC FACTOR procedure in SAS (v9.4; SAS Institute Inc., Cary, NC USA). This procedure was run with prespecified numbers of factors (2-8) to determine which value best explained the variation in the current sample’s FV intakes. The 6-factor set was chosen based on scree plot readings, eigenvalues, total variance explained and meaningful interpretation of the individual factor loadings. Background and discussion on these methods have been described ^89^.

### Measures of the Metabolites

Metabolic profiling of plasma samples was performed by Metabolon, Inc (Morrisville, NC USA) as described for this population ^90^. Briefly, frozen plasma samples were shipped on dry ice, and stored at −80°C until analysis. After methanol extraction of proteins, metabolomic analysis employed ultrahigh-performance liquid chromatography-tandem mass spectroscopy. Individual metabolites were identified by referring to a library of over 4500 purified standards for retention time/index, mass-to-charge ratio, and chromatographic data, and then quantified by estimating the AUC of the peaks. Metabolites were log-transformed for entry into analysis. The median relative standard deviation for internal standards (a measure of instrument variability akin to coefficient of variation) was 5%. After normalization across samples, 525 metabolites passed quality control. See section “Additional Covariates,” below, for details on other measured outcomes. Metabolite abbreviations are defined in **Table S9**.

### Additional Covariates

Data on sex, age, menopausal status, estrogen use, and osteoporosis medication use were obtained through questionnaire during BPROS visits. Hormone status of women and sex of participants were used to derive an estrogenic status variable (male, estrogenic female, non-estrogenic female). Height and weight were measured in duplicate per standard methods. BMI was calculated as weight (kg)/height (m)^2^. A fasting blood sample was collected by a bilingual certified phlebotomist to assess plasma 25-hydroxyvitamin D (25OHD). Blood samples were collected in evacuated EDTA tubes and centrifuged to separate plasma. A ^125^I radioimmunoassay (DiaSorin, Inc., Stillwater, MN, USA; manufacturer procedures: 68100E) ^91^ was used to obtain plasma 25OHD measures; the intra-assay coefficient of variation was 10.8% and inter-assay coefficient of variation was 9.4%. All other covariates, including educational attainment (<8^th^ grade, 9^th^-12^th^ grade and some college or higher), alcohol consumption, smoking status, physical activity, total dietary calcium intake and total energy intake were obtained by questionnaires during the 2-yr follow-up visit ^78^. The physical activity index (PAI) is a variation of the Framingham PAI, a weighted 24-h score of typical daily activity, based on hours spent doing heavy, moderate, light, or sedentary activity plus sleeping ^92^. Dietary calcium intake (mg) and total energy intake (kcal) were estimated from the FFQ ^83^.

### Bioinformatics and Statistical Analysis

#### Associations of plasma metabolites with OS status and FV intakes

Differences in plasma metabolite concentrations between disease groups (OS vs no-OS) were compared using logistic regression while adjusted for age, smoking, alcohol use, height, and physical activity. Metabolites were considered nominally significantly different between groups at P≤0.05. Associations between plasma metabolites and total FV intake were assessed with linear regression while controlling for age, smoking, alcohol use, physical activity, education, and total energy intake. Associations were considered nominally significant at P≤0.05. A positive beta coefficient indicates higher metabolite concentration in those with higher intake of FV as combined servings per day. For OS status, a positive beta coefficient indicates increased metabolite concentration in those with OS, and correspondingly a negative beta coefficient indicates lower concentration in those with OS.

#### Identification of enzyme inhibitors related to metabolites significantly associated with OS status or FV intake

Enzyme inhibitor data were mined from BRENDA to identify metabolites with published data indicating inhibitory action on a given Enzyme Commission (EC) number ^93^. EC number-pathway assignments were taken from KEGG. Lower inhibitory action (negative beta) is interpreted as supporting reduced inhibition of that particular enzyme.

#### Pathway analysis

Metabolites significantly associated with OS status or with total FV intake were analyzed for enrichment in various biological pathways, functional modules, bioprocesses and diseases. Software included Mbrole 2.0 ^94^, MetaboAnalyst ^95^, and Reactome ^96^, with analyses run using default parameters, including FDR correction of P-values. These analysis packages support assignment of a metabolite to several, often overlapping, functional pathways and modules. Data were analyzed as women and men separately. Biological interpretation of the resulting enriched functional and pathway sets was conducted for those sets represented by two or more significantly different metabolites.

#### Correlation of OS-associated metabolites with intake of specific FV food groupings

Correlation coefficients were calculated as Spearman’s rho rank correlations, followed by hierarchical clustering of the resulting correlation coefficients using the top six PCA factors for intake of FV and sex-specific sets of OS-associated metabolites. This work was performed in R (v 3.5.1; R Core Team (2018) R: A language and environment for statistical computing. R Foundation for Statistical Computing, Vienna, Austria. URL: https://www.R-project.org/) and RStudio (v 1.1.456) with the `tidyverse`, `ggpubr`, `Hmisc` (type=“spearman”)`,reshape2` and `stats` packages. We used the `stats::hclust` function for hierarchical clustering. To identify metabolite groups illustrative of bone health and OS within the disease-associated metabolite profiles, PCA of the osteoporosis-associated metabolites was performed, in R with the `factoextra` library and the `stats::prcomp` function (center=“TRUE”, scale.=“TRUE”), in addition to the above libraries and functions. Significance was evaluated with the `Hmisc::rcorr` and `t.test` functions, with P<0.05 considered significant. The `glm` function (family=“binomial”) was used to test logit models for predicting OS status. Molecules of interest were queried in the scientific literature for evidence relating to OS, bone health, or osteoblast or osteoclast function.

All statistical tests were two-sided and considered statistically significant at p<0.05.

## Data Availability

The datasets generated during and/or analyzed during the current study are available from the corresponding author on reasonable request.

## Abbreviations

AUC: area under the curve
BCAA: branched-chain amino acid
BPRHS: Boston Puerto Rican Health Study
BPROS: Boston Puerto Rican Osteoporosis Study
DHA: docosahexaenoic acid
DPA: docosahexaenoic acid
EC: Enzyme Commission
EPA: eicosapentaenoic acid
FDR: false discovery rate
FFQ: food frequency questionnaire
FV: fruit and vegetable
KEGG: Kyoto Encyclopedia of Genes and Genomes
NSAID: nonsteroidal anti-inflammatory drug
OS: osteoporosis
PCA: principal components analysis
PUFA: poly-unsaturated fatty acids
USDA: United States Department of Agriculture

## Acknowledgements

The Boston Puerto Rican Health Study and Osteoporosis Study have been supported by NIH P50 HL105185, P01 AG023394, R01 AG055948, R01 AG027087 and R01 AR072741. Grant 201806105018 from the US Department of Agriculture, under agreement no. 8050-51000-098-00D partially supported this work, as well as support for SEN’s time by NIAMS-KO1AR067894. Any opinions, findings, conclusion, or recommendations expressed in this publication are those of the authors and do not necessarily reflect the view of the US Department of Agriculture. Mention of trade names or commercial products in this publication is solely for providing specific information and does not imply recommendation or endorsement by the U.S. Department of Agriculture. The USDA is an equal opportunity provider and employer.

## Author Contributions

Study design: KLT, KMM, LDP and SEN. Study conduct: KLT. Data collection: SEN and BDH. Data analysis: LDP, CQL and KMM. Data interpretation: LDP, CQL, KMM, KLT, SEN, and JJC. Drafting manuscript: LDP and KMM. Revising manuscript content: KMM, LDP, SEN, KLT, JJC, CQL, JMO, and BDH. All authors take responsibility for approving the final version of manuscript and for the integrity of the data analysis.

## Code Availability Statement

All computer code used to generate data within this manuscript are available from the corresponding authors upon reasonable request.

## Figure legends

**Figure S1.**
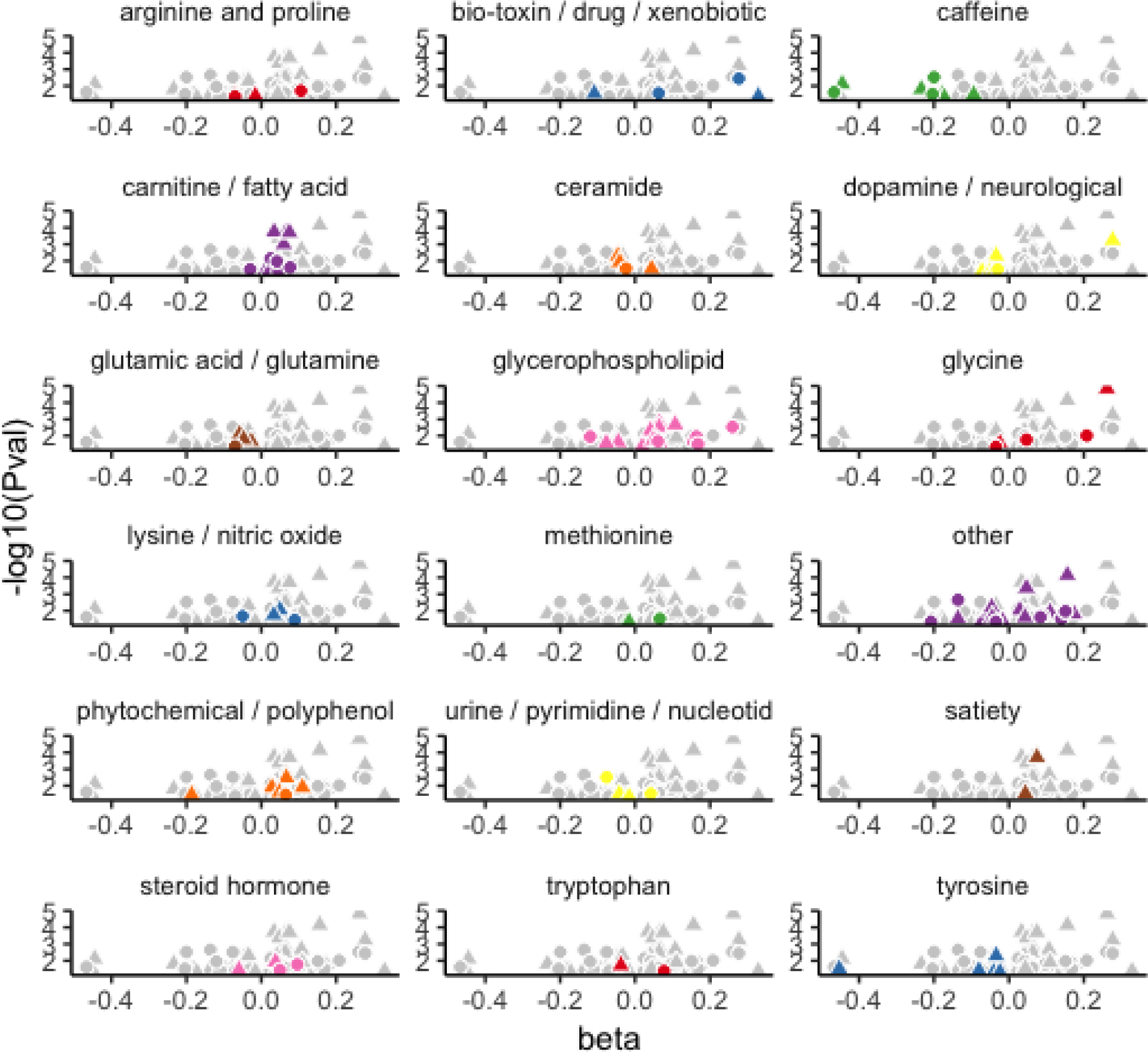
Plasma metabolites significantly associated with FV intake. Each panel shows a plot of beta *vs* -log_10_(P-value) for all metabolites significantly associated with OS (gray) and a specific metabolite class highlighted in color. Data for women are plotted with triangles, for men with circles, and are derived from **Table S1**, which lists 66 metabolite-FV associations for women and 38 for men.

**Figure S2.**
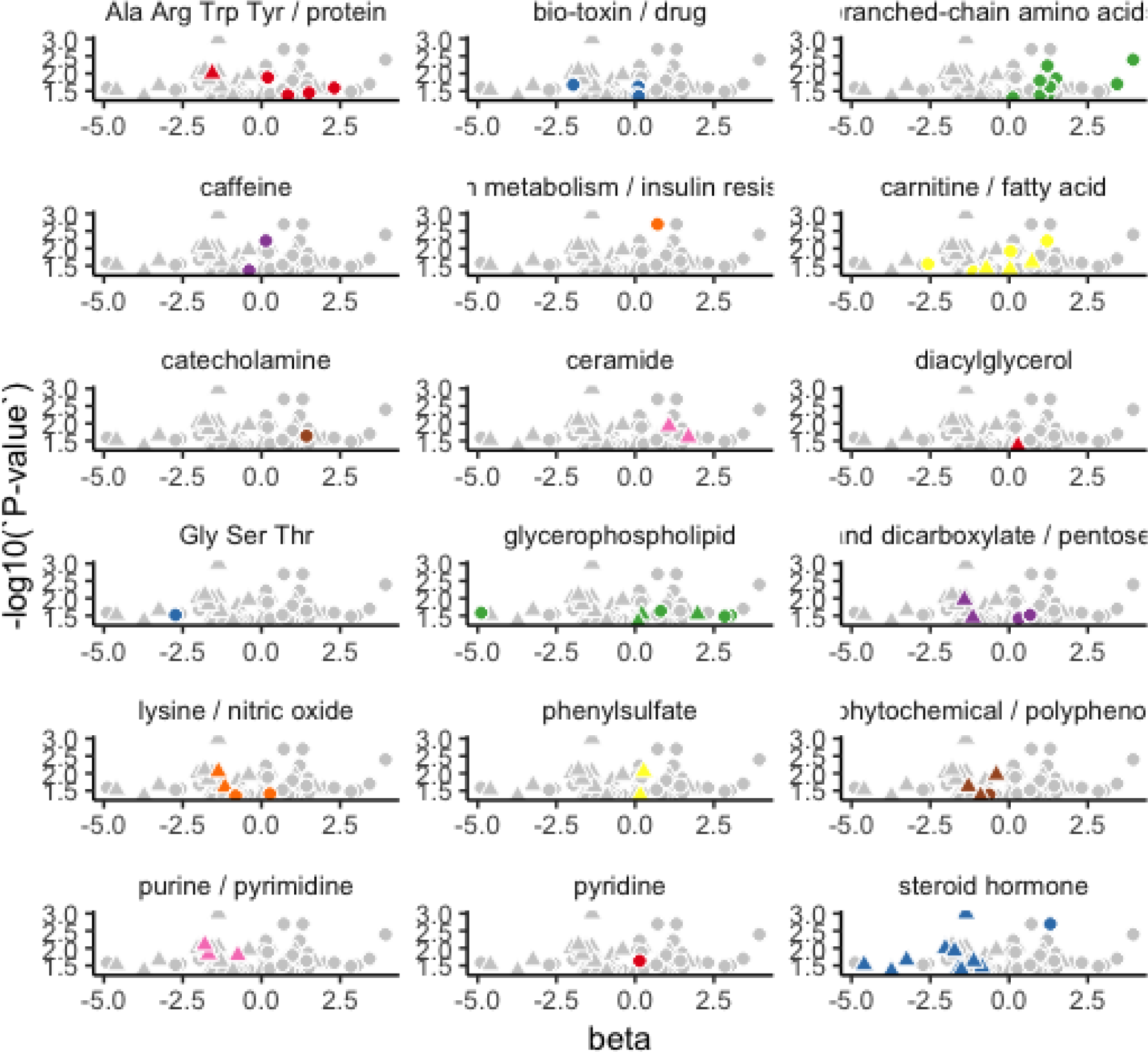
Plasma metabolites significantly associated with OS status. Each panel depicts a volcano plot, beta *vs* -log_10_(P-value), for all metabolites significantly associated with OS (gray) and a distinct metabolite class highlighted in color. Data for women are plotted with triangles, for men with circles, and are derived from **Table S5**, which lists 33 metabolite-FV associations for women and 40 for men.

**Figure S3.**
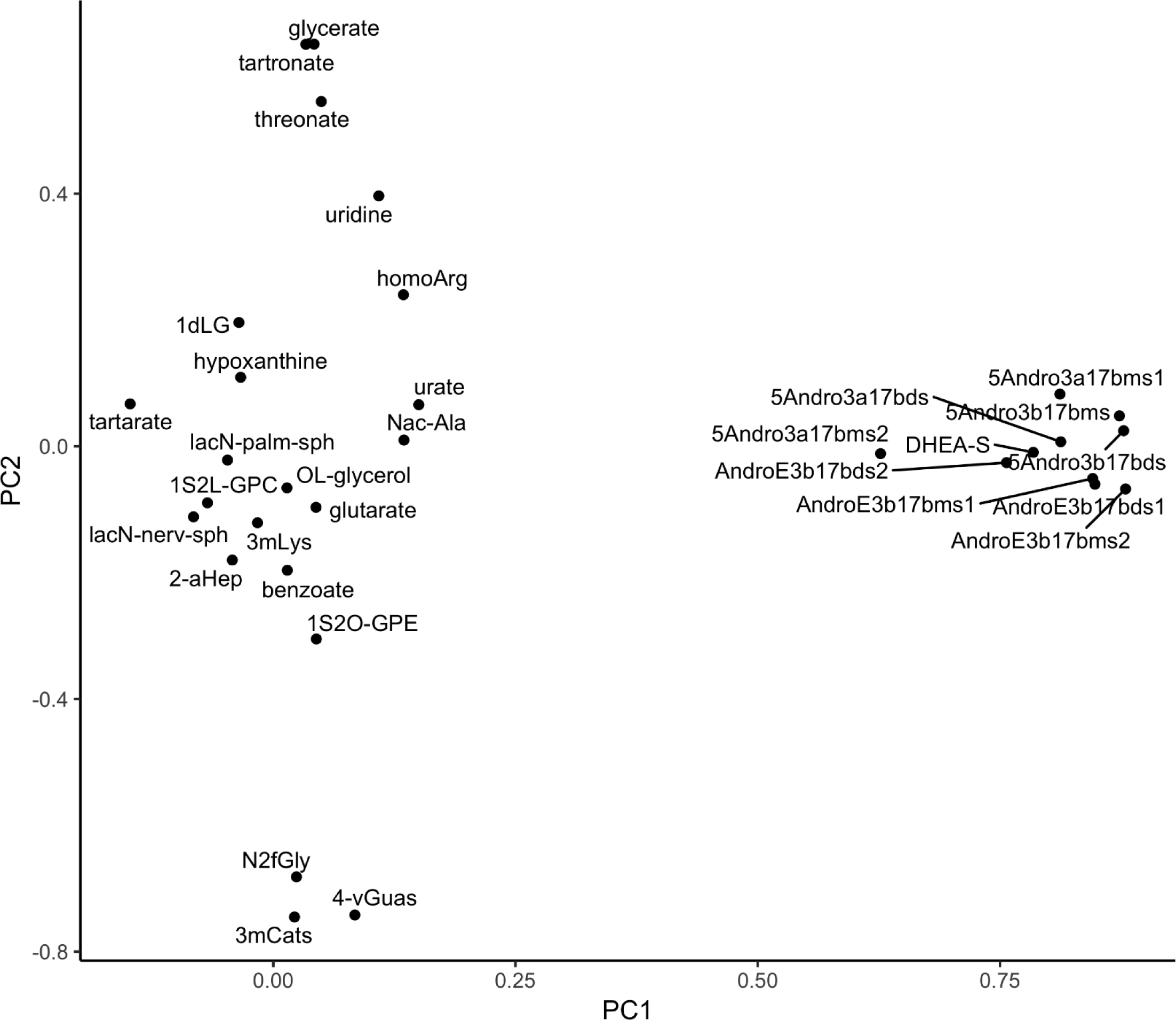
PCA of OS-associated metabolites in BPROS women. OS-associated metabolites are plotted by factor 1 (PC1) vs factor 2 (PC2), which in turn represent 21.2% and 10.0% of the total variance. The group of ten steroid hormone metabolites all have highly positive PC1 values and form a tight cluster. Abbreviations for metabolites are defined in **Table S9**.

**Figure S4.**
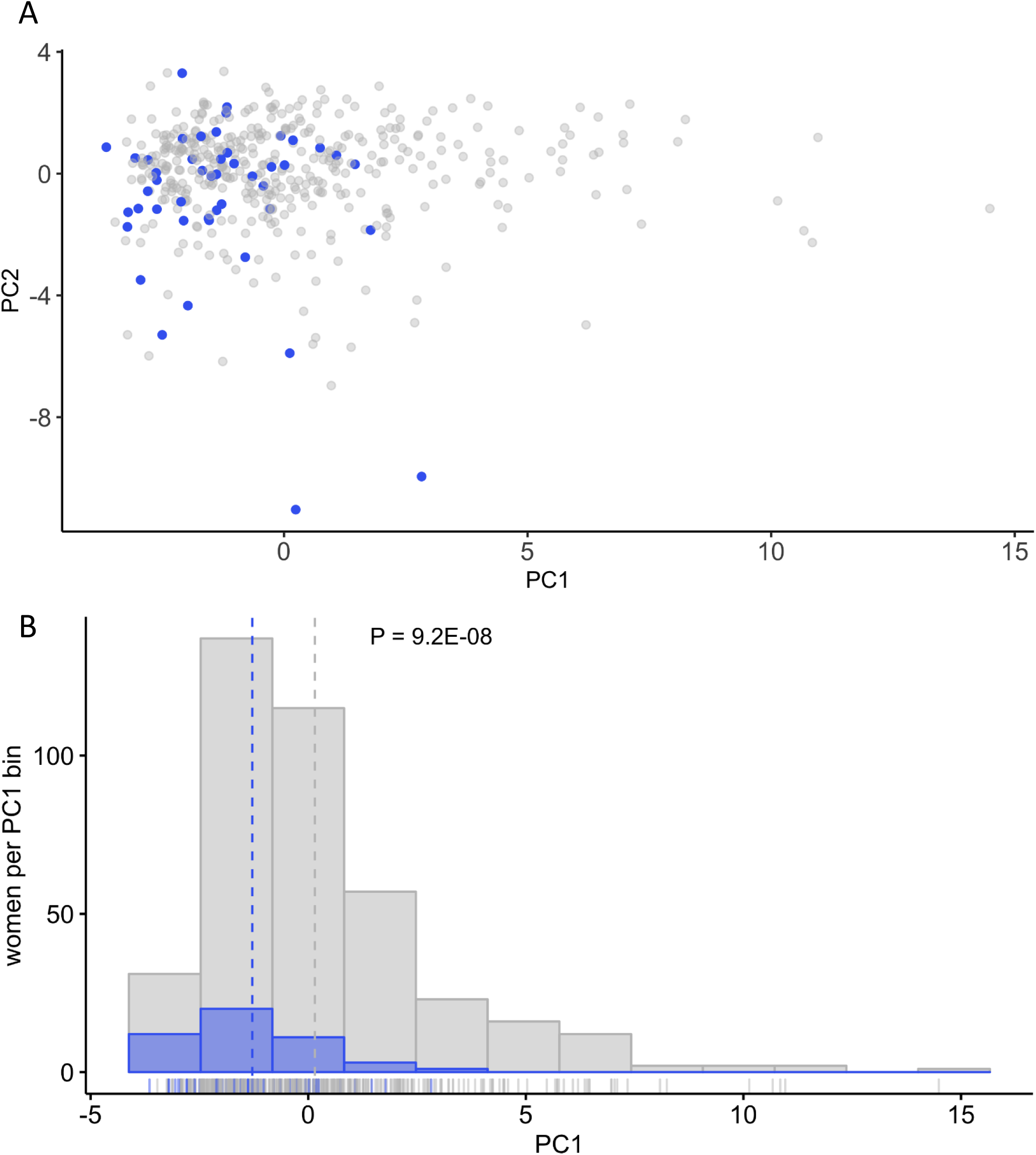
PC1 values are a strong indicator of OS status in BPROS women. A) Female participants are plotted according to PC1 and PC2 values with OS status depicted as gray without and blue with osteoporosis. B) A histogram of 12 evenly spaced bins of PC1 values indicates that the mean PC1 values (dotted line) and the distribution of all values for women with OS are shifted toward lower PC1 values than for women without disease (P=9.2E-08). Running along the x-axis is a density plot of all women based on their PC1 values, colored by OS status.

**Figure S5.**
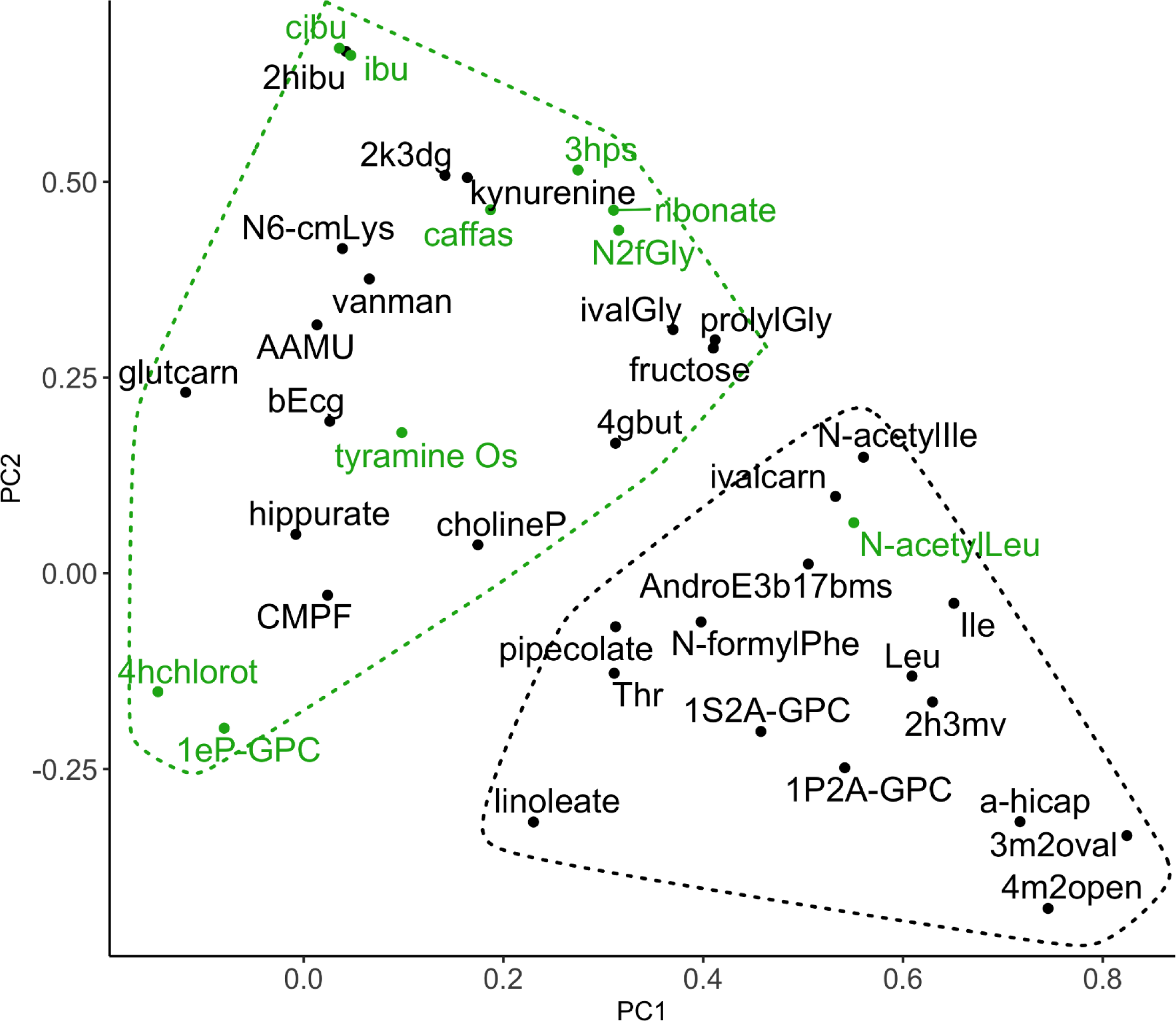
PCA of OS-associated metabolites in BPROS men. OS-associated metabolites are plotted by factor 1 (PC1) vs factor 2 (PC2), which represent 15.1% and 10.9% of the total variance, respectively. Green data points are those metabolites whose correlations with FV factor 6 (dark leafy greens, all P<0.10) are exactly opposite to the association with OS incidence, per **Figure 3B**. Abbreviations for metabolites are defined in **Table S9**.

## References

1 Noel, S. E. et al. Prevalence of Osteoporosis and Low Bone Mass Among Puerto Rican Older Adults. J Bone Miner Res, doi:10.1002/jbmr.3315 (2017).

2 Alswat, K. A. Gender Disparities in Osteoporosis. Journal of Clinical Medicine Research 9, 382–387, doi:10.14740/jocmr2970w (2017).

3 Cauley, J. A. Public health impact of osteoporosis. J Gerontol A Biol Sci Med Sci 68, 1243–1251, doi:10.1093/gerona/glt093 (2013).

4 Magaziner, J. et al. Recovery from hip fracture in eight areas of function. J Gerontol A Biol Sci Med Sci 55, M498–507 (2000).

5 Marks, R., Allegrante, J. P., Ronald MacKenzie, C. & Lane, J. M. Hip fractures among the elderly: causes, consequences and control. Ageing Res Rev 2, 57–93 (2003).

6 Imaz, I. et al. Poor bisphosphonate adherence for treatment of osteoporosis increases fracture risk: systematic review and meta-analysis. Osteoporos Int 21, 1943–1951, doi:10.1007/s00198-009-1134-4 (2010).

7 McCombs, J. S., Thiebaud, P., McLaughlin-Miley, C. & Shi, J. Compliance with drug therapies for the treatment and prevention of osteoporosis. Maturitas 48, 271–287, doi:10.1016/j.maturitas.2004.02.005 (2004).

8 Weycker, D., Macarios, D., Edelsberg, J. & Oster, G. Compliance with drug therapy for postmenopausal osteoporosis. Osteoporos Int 17, 1645–1652, doi:10.1007/s00198-006-0179-x (2006).

9 Morgan, K. T. Nutritional determinants of bone health. J Nutr Elder 27, 3–27, doi:10.1080/01639360802059670 (2008).

10 Tucker, K. L. Osteoporosis prevention and nutrition. Curr Osteoporos Rep 7, 111–117 (2009).

11 Chen, Y. M., Ho, S. C. & Woo, J. L. Greater fruit and vegetable intake is associated with increased bone mass among postmenopausal Chinese women. Br J Nutr 96, 745–751 (2006).

12 Prynne, C. J. et al. Fruit and vegetable intakes and bone mineral status: a cross sectional study in 5 age and sex cohorts. Am J Clin Nutr 83, 1420–1428 (2006).

13 Qiu, R. et al. Greater Intake of Fruit and Vegetables Is Associated with Greater Bone Mineral Density and Lower Osteoporosis Risk in Middle-Aged and Elderly Adults. PLoS One 12, e0168906, doi:10.1371/journal.pone.0168906 (2017).

14 Tucker, K. L. et al. Potassium, magnesium, and fruit and vegetable intakes are associated with greater bone mineral density in elderly men and women. Am J Clin Nutr 69, 727–736 (1999).

15 Byberg, L., Bellavia, A., Orsini, N., Wolk, A. & Michaelsson, K. Fruit and vegetable intake and risk of hip fracture: a cohort study of Swedish men and women. J Bone Miner Res 30, 976–984, doi:10.1002/jbmr.2384 (2015).

16 Macdonald, H. M., New, S. A., Golden, M. H., Campbell, M. K. & Reid, D. M. Nutritional associations with bone loss during the menopausal transition: evidence of a beneficial effect of calcium, alcohol, and fruit and vegetable nutrients and of a detrimental effect of fatty acids. Am J Clin Nutr 79, 155–165, doi:10.1093/ajcn/79.1.155 (2004).

17 Kaptoge, S. et al. Effects of dietary nutrients and food groups on bone loss from the proximal femur in men and women in the 7th and 8th decades of age. Osteoporos Int 14, 418–428, doi:10.1007/s00198-003-1391-6 (2003).

18 Benetou, V. et al. Fruit and Vegetable Intake and Hip Fracture Incidence in Older Men and Women: The CHANCES Project. J Bone Miner Res 31, 1743–1752, doi:10.1002/jbmr.2850 (2016).

19 Macdonald, H. M. et al. Effect of potassium citrate supplementation or increased fruit and vegetable intake on bone metabolism in healthy postmenopausal women: a randomized controlled trial. Am J Clin Nutr 88, 465–474, doi:10.1093/ajcn/88.2.465 (2008).

20 Neville, C. E. et al. Effect of increased fruit and vegetable consumption on bone turnover in older adults: a randomised controlled trial. Osteoporos Int 25, 223–233, doi:10.1007/s00198-013-2402-x (2014).

21 Nieves, J. W. Skeletal effects of nutrients and nutraceuticals, beyond calcium and vitamin D. Osteoporos Int 24, 771–786, doi:10.1007/s00198-012-2214-4 (2013).

22 Weaver, C. M., Alekel, D. L., Ward, W. E. & Ronis, M. J. Flavonoid intake and bone health. J Nutr Gerontol Geriatr 31, 239–253, doi:10.1080/21551197.2012.698220 (2012).

23 Hansen, H. S. Role of anorectic N-acylethanolamines in intestinal physiology and satiety control with respect to dietary fat. Pharmacol Res 86, 18–25, doi:10.1016/j.phrs.2014.03.006 (2014).

24 Pettersen, J. E. & Jellum, E. The identification and metabolic origin of 2-furoylglycine and 2,5-furandicarboxylic acid in human urine. Clin Chim Acta 41, 199–207, doi:10.1016/0009-8981(72)90512-8 (1972).

25 Luo, S. et al. Increased intake of vegetables, but not fruits, may be associated with reduced risk of hip fracture: A meta-analysis. Sci Rep 6, 19783, doi:10.1038/srep19783 (2016).

26 Sahni, S., Mangano, K. M., McLean, R. R., Hannan, M. T. & Kiel, D. P. Dietary Approaches for Bone Health: Lessons from the Framingham Osteoporosis Study. Curr Osteoporos Rep 13, 245–255, doi:10.1007/s11914-015-0272-1 (2015).

27 Klinder, A. et al. Impact of increasing fruit and vegetables and flavonoid intake on the human gut microbiota. Food Funct 7, 1788–1796, doi:10.1039/c5fo01096a (2016).

28 Trost, K. et al. Host: Microbiome co-metabolic processing of dietary polyphenols - An acute, single blinded, cross-over study with different doses of apple polyphenols in healthy subjects. Food Res Int 112, 108–128, doi:10.1016/j.foodres.2018.06.016 (2018).

29 Kelly, O. J., Gilman, J. C., Kim, Y. & Ilich, J. Z. Long-chain polyunsaturated fatty acids may mutually benefit both obesity and osteoporosis. Nutr Res 33, 521–533, doi:10.1016/j.nutres.2013.04.012 (2013).

30 Boeyens, J. C. et al. Effects of omega3- and omega6-polyunsaturated fatty acids on RANKL-induced osteoclast differentiation of RAW264.7 cells: a comparative in vitro study. Nutrients 6, 2584–2601, doi:10.3390/nu6072584 (2014).

31 Pilz, S. et al. Associations of homoarginine with bone metabolism and density, muscle strength and mortality: cross-sectional and prospective data from 506 female nursing home patients. Osteoporos Int 24, 377–381, doi:10.1007/s00198-012-1950-9 (2013).

32 Deepak, V., Kruger, M. C., Joubert, A. & Coetzee, M. Piperine alleviates osteoclast formation through the p38/c-Fos/NFATc1 signaling axis. Biofactors 41, 403–413, doi:10.1002/biof.1241 (2015).

33 Refaey, M. E. et al. Kynurenine, a Tryptophan Metabolite That Accumulates With Age, Induces Bone Loss. J Bone Miner Res 32, 2182–2193, doi:10.1002/jbmr.3224 (2017).

34 Jovanovich, A. et al. Deoxycholic Acid, a Metabolite of Circulating Bile Acids, and Coronary Artery Vascular Calcification in CKD. Am J Kidney Dis 71, 27–34, doi:10.1053/j.ajkd.2017.06.017 (2018).

35 Poloni, S. et al. Leptin concentrations and SCD-1 indices in classical homocystinuria: Evidence for the role of sulfur amino acids in the regulation of lipid metabolism. Clin Chim Acta 473, 82–88, doi:10.1016/j.cca.2017.08.005 (2017).

36 Gundberg, C. M., Lian, J. B., Gallop, P. M. & Steinberg, J. J. Urinary gamma-carboxyglutamic acid and serum osteocalcin as bone markers: studies in osteoporosis and Paget’s disease. J Clin Endocrinol Metab 57, 1221–1225, doi:10.1210/jcem-57-6-1221 (1983).

37 Baek, J. M. et al. Nicotinamide phosphoribosyltransferase inhibits receptor activator of nuclear factor-kappaB ligand-induced osteoclast differentiation in vitro. Mol Med Rep 15, 784–792, doi:10.3892/mmr.2016.6069 (2017).

38 Phang, J. M., Liu, W. & Zabirnyk, O. Proline metabolism and microenvironmental stress. Annu Rev Nutr 30, 441–463, doi:10.1146/annurev.nutr.012809.104638 (2010).

39 Oyen, J. et al. Plasma dimethylglycine, nicotine exposure and risk of low bone mineral density and hip fracture: the Hordaland Health Study. Osteoporos Int 26, 1573–1583, doi:10.1007/s00198-015-3030-4 (2015).

40 Lopez-Gonzalez, A. A. et al. Phytate (myo-inositol hexaphosphate) and risk factors for osteoporosis. J Med Food 11, 747–752, doi:10.1089/jmf.2008.0087 (2008).

41 Akiyama, M., Nakahama, K. & Morita, I. Impact of docosahexaenoic acid on gene expression during osteoclastogenesis in vitro--a comprehensive analysis. Nutrients 5, 3151–3162, doi:10.3390/nu5083151 (2013).

42 Yeung, D. K. et al. Osteoporosis is associated with increased marrow fat content and decreased marrow fat unsaturation: a proton MR spectroscopy study. J Magn Reson Imaging 22, 279–285, doi:10.1002/jmri.20367 (2005).

43 Patsch, J. M. et al. Bone marrow fat composition as a novel imaging biomarker in postmenopausal women with prevalent fragility fractures. J Bone Miner Res 28, 1721–1728, doi:10.1002/jbmr.1950 (2013).

44 Gao, B. et al. Dose-response estrogen promotes osteogenic differentiation via GPR40 (FFAR1) in murine BMMSCs. Biochimie 110, 36–44, doi:10.1016/j.biochi.2015.01.001 (2015).

45 Candow, D. G., Chilibeck, P. D. & Forbes, S. C. Creatine supplementation and aging musculoskeletal health. Endocrine 45, 354–361, doi:10.1007/s12020-013-0070-4 (2014).

46 Bechtel, Y. C. et al. Caffeine metabolism before and after liver transplantation. Int J Clin Pharmacol Ther 39, 53–60, doi:10.5414/cpp39053 (2001).

47 Bhatta, A. et al. Deregulation of arginase induces bone complications in high-fat/high-sucrose diet diabetic mouse model. Mol Cell Endocrinol 422, 211–220, doi:10.1016/j.mce.2015.12.005 (2016).

48 Conigrave, A. D., Brown, E. M. & Rizzoli, R. Dietary protein and bone health: roles of amino acid-sensing receptors in the control of calcium metabolism and bone homeostasis. Annu Rev Nutr 28, 131–155, doi:10.1146/annurev.nutr.28.061807.155328 (2008).

49 Manolagas, S. C., Kousteni, S. & Jilka, R. L. Sex steroids and bone. Recent Prog Horm Res 57, 385–409 (2002).

50 Syed, F. & Khosla, S. Mechanisms of sex steroid effects on bone. Biochem Biophys Res Commun 328, 688–696, doi:10.1016/j.bbrc.2004.11.097 (2005).

51 Imai, Y. et al. Estrogens maintain bone mass by regulating expression of genes controlling function and life span in mature osteoclasts. Ann N Y Acad Sci 1173 Suppl 1, E31–39, doi:10.1111/j.1749-6632.2009.04954.x (2009).

52 Eisenberger, M. A. et al. Phase I and clinical pharmacology of a type I and II, 5-alpha-reductase inhibitor (LY320236) in prostate cancer: elevation of estradiol as possible mechanism of action. Urology 63, 114–119, doi:10.1016/j.urology.2003.08.017 (2004).

53 Amory, J. K. et al. The effect of 5alpha-reductase inhibition with dutasteride and finasteride on bone mineral density, serum lipoproteins, hemoglobin, prostate specific antigen and sexual function in healthy young men. J Urol 179, 2333–2338, doi:10.1016/j.juro.2008.01.145 (2008).

54 Borst, S. E. et al. Anabolic effects of testosterone are preserved during inhibition of 5alpha-reductase. Am J Physiol Endocrinol Metab 293, E507–514, doi:10.1152/ajpendo.00130.2007 (2007).

55 Matsumoto, A. M. et al. The long-term effect of specific type II 5alpha-reductase inhibition with finasteride on bone mineral density in men: results of a 4-year placebo controlled trial. J Urol 167, 2105–2108 (2002).

56 Jacobsen, S. J., Cheetham, T. C., Haque, R., Shi, J. M. & Loo, R. K. Association between 5-alpha reductase inhibition and risk of hip fracture. JAMA 300, 1660–1664, doi:10.1001/jama.300.14.1660 (2008).

57 Downey, P. A. & Siegel, M. I. Bone biology and the clinical implications for osteoporosis. Phys Ther 86, 77–91, doi:10.1093/ptj/86.1.77 (2006).

58 Heshmati, H. M. et al. Role of low levels of endogenous estrogen in regulation of bone resorption in late postmenopausal women. J Bone Miner Res 17, 172–178, doi:10.1359/jbmr.2002.17.1.172 (2002).

59 Reed, M. J., Purohit, A., Woo, L. W., Newman, S. P. & Potter, B. V. Steroid sulfatase: molecular biology, regulation, and inhibition. Endocr Rev 26, 171–202, doi:10.1210/er.2004-0003 (2005).

60 Dias, N. J. & Selcer, K. W. Steroid sulfatase in the human MG-63 preosteoblastic cell line: Antagonistic regulation by glucocorticoids and NFkappaB. Mol Cell Endocrinol 420, 85–96, doi:10.1016/j.mce.2015.11.029 (2016).

61 Klein, B. Y., Kerem, Z. & Rojansky, N. LDL induces Saos2 osteoblasts death via Akt pathways responsive to a neutral sphingomyelinase inhibitor. J Cell Biochem 98, 661–671, doi:10.1002/jcb.20807 (2006).

62 He, J. H., Tong, N. W., Li, H. Q. & Wu, J. [Effects of L-threonate on bone resorption by osteoclasts in vitro]. Sichuan Da Xue Xue Bao Yi Xue Ban 36, 225–228 (2005).

63 Rowe, D. J., Ko, S., Tom, X. M., Silverstein, S. J. & Richards, D. W. Enhanced production of mineralized nodules and collagenous proteins in vitro by calcium ascorbate supplemented with vitamin C metabolites. J Periodontol 70, 992–999, doi:10.1902/jop.1999.70.9.992 (1999).

64 Ohtsuki, S. et al. Mouse reduced in osteosclerosis transporter functions as an organic anion transporter 3 and is localized at abluminal membrane of blood-brain barrier. J Pharmacol Exp Ther 309, 1273–1281, doi:10.1124/jpet.103.063370 (2004).

65 Chen, J. R. et al. Diet-derived phenolic acids regulate osteoblast and adipocyte lineage commitment and differentiation in young mice. J Bone Miner Res 29, 1043–1053, doi:10.1002/jbmr.2034 (2014).

66 Hu, J. R. et al. Serum metabolites are associated with all-cause mortality in chronic kidney disease. Kidney Int 94, 381–389, doi:10.1016/j.kint.2018.03.008 (2018).

67 Kalaska, B. et al. Elevated Levels of Peripheral Kynurenine Decrease Bone Strength in Rats with Chronic Kidney Disease. Front Physiol 8, 836, doi:10.3389/fphys.2017.00836 (2017).

68 Ehrlich, H. et al. Modification of collagen in vitro with respect to formation of Nepsilon-carboxymethyllysine. Int J Biol Macromol 44, 51–56, doi:10.1016/j.ijbiomac.2008.10.001 (2009).

69 Hein, G., Wiegand, R., Lehmann, G., Stein, G. & Franke, S. Advanced glycation end-products pentosidine and N epsilon-carboxymethyllysine are elevated in serum of patients with osteoporosis. Rheumatology (Oxford) 42, 1242–1246, doi:10.1093/rheumatology/keg324 (2003).

70 Adams, S. B., Jr. et al. Global metabolic profiling of human osteoarthritic synovium. Osteoarthritis Cartilage 20, 64–67, doi:10.1016/j.joca.2011.10.010 (2012).

71 Zhai, G. et al. Serum branched-chain amino acid to histidine ratio: a novel metabolomic biomarker of knee osteoarthritis. Ann Rheum Dis 69, 1227–1231, doi:10.1136/ard.2009.120857 (2010).

72 Agriculture, U. S. D. o. H. a. H. S. a. U. S. D. o. (December 2015).

73 Booth, S. L. et al. Dietary vitamin K intakes are associated with hip fracture but not with bone mineral density in elderly men and women. Am J Clin Nutr 71, 1201–1208, doi:10.1093/ajcn/71.5.1201 (2000).

74 Feskanich, D. et al. Vitamin K intake and hip fractures in women: a prospective study. Am J Clin Nutr 69, 74–79, doi:10.1093/ajcn/69.1.74 (1999).

75 Noel, S. et al. Risk Factors for Bone Health in Older Puerto Rican Adults (OR18-05-19). Curr Dev Nutr 3, doi:10.1093/cdn/nzz028.OR18-05-19 (2019).

76 Sahni, S. et al. Protective effect of total and supplemental vitamin C intake on the risk of hip fracture--a 17-year follow-up from the Framingham Osteoporosis Study. Osteoporos Int 20, 1853–1861, doi:10.1007/s00198-009-0897-y (2009).

77 Sahni, S. et al. Inverse association of carotenoid intakes with 4-y change in bone mineral density in elderly men and women: the Framingham Osteoporosis Study. Am J Clin Nutr 89, 416–424, doi:10.3945/ajcn.2008.26388 (2009).

78 Tucker, K. L. et al. The Boston Puerto Rican Health Study, a longitudinal cohort study on health disparities in Puerto Rican adults: challenges and opportunities. BMC Public Health 10, 107, doi:10.1186/1471-2458-10-107 (2010).

79 Harris, P. A. et al. The REDCap consortium: Building an international community of software platform partners. J Biomed Inform 95, 103208, doi:10.1016/j.jbi.2019.103208 (2019).

80 Harris, P. A. et al. Research electronic data capture (REDCap)--a metadata-driven methodology and workflow process for providing translational research informatics support. J Biomed Inform 42, 377–381, doi:10.1016/j.jbi.2008.08.010 (2009).

81 White, J., Harris, S. S., Dallal, G. E. & Dawson-Hughes, B. Precision of single vs bilateral hip bone mineral density scans. J Clin Densitom 6, 159–162 (2003).

82 Kanis, J. A. et al. A reference standard for the description of osteoporosis. Bone 42, 467–475, doi:10.1016/j.bone.2007.11.001 (2008).

83 Tucker, K. L., Bianchi, L. A., Maras, J. & Bermudez, O. I. Adaptation of a food frequency questionnaire to assess diets of Puerto Rican and non-Hispanic adults. Am J Epidemiol 148, 507–518, doi:10.1093/oxfordjournals.aje.a009676 (1998).

84 Bermudez, O. I., Ribaya-Mercado, J. D., Talegawkar, S. A. & Tucker, K. L. Hispanic and non-Hispanic white elders from Massachusetts have different patterns of carotenoid intake and plasma concentrations. J Nutr 135, 1496–1502, doi:10.1093/jn/135.6.1496 (2005).

85 Gao, X., Martin, A., Lin, H., Bermudez, O. I. & Tucker, K. L. alpha-Tocopherol intake and plasma concentration of Hispanic and non-Hispanic white elders is associated with dietary intake pattern. J Nutr 136, 2574–2579, doi:10.1093/jn/136.10.2574 (2006).

86 Ye, X., Maras, J. E., Bakun, P. J. & Tucker, K. L. Dietary intake of vitamin B-6, plasma pyridoxal 5’-phosphate, and homocysteine in Puerto Rican adults. J Am Diet Assoc 110, 1660–1668, doi:10.1016/j.jada.2010.08.006 (2010).

87 Kwan, L. L., Bermudez, O. I. & Tucker, K. L. Low vitamin B-12 intake and status are more prevalent in Hispanic older adults of Caribbean origin than in neighborhood-matched non-Hispanic whites. J Nutr 132, 2059–2064, doi:10.1093/jn/132.7.2059 (2002).

88 Bhupathiraju, S. N. & Tucker, K. L. Greater variety in fruit and vegetable intake is associated with lower inflammation in Puerto Rican adults. Am J Clin Nutr 93, 37–46, doi:10.3945/ajcn.2010.29913 (2011).

89 Newby, P. K. & Tucker, K. L. Empirically derived eating patterns using factor or cluster analysis: a review. Nutr Rev 62, 177–203, doi:10.1301/nr.2004.may.177-203 (2004).

90 Lai, C. Q. et al. Epigenomics and metabolomics reveal the mechanism of the APOA2-saturated fat intake interaction affecting obesity. Am J Clin Nutr 108, 188–200, doi:10.1093/ajcn/nqy081 (2018).

91 Wang, S. et al. Fermented milk supplemented with probiotics and prebiotics can effectively alter the intestinal microbiota and immunity of host animals. J Dairy Sci 95, 4813–4822, doi:10.3168/jds.2012-5426 (2012).

92 Kannel, W. B. & Sorlie, P. Some health benefits of physical activity. The Framingham Study. Arch Intern Med 139, 857–861 (1979).

93 Schomburg, I. et al. The BRENDA enzyme information system-From a database to an expert system. J Biotechnol 261, 194–206, doi:10.1016/j.jbiotec.2017.04.020 (2017).

94 Lopez-Ibanez, J., Pazos, F. & Chagoyen, M. MBROLE 2.0-functional enrichment of chemical compounds. Nucleic Acids Res 44, W201–204, doi:10.1093/nar/gkw253 (2016).

95 Chong, J. et al. MetaboAnalyst 4.0: towards more transparent and integrative metabolomics analysis. Nucleic Acids Res 46, W486–W494, doi:10.1093/nar/gky310 (2018).

96 Fabregat, A. et al. The Reactome pathway Knowledgebase. Nucleic Acids Res 44, D481–487, doi:10.1093/nar/gkv1351 (2016).

